# Global COVID-19 transmission rate is influenced by precipitation seasonality and the speed of climate temperature warming

**DOI:** 10.1101/2020.04.10.20060459

**Authors:** Katsumi Chiyomaru, Kazuhiro Takemoto

**Author notes:** Corresponding author’s.

## Abstract

The novel coronavirus disease 2019 (COVID-19) became a rapidly spreading worldwide epidemic; thus, it is a global priority to reduce the speed of the epidemic spreading. Several studies predicted that high temperature and humidity could reduce COVID-19 transmission. However, exceptions exist to this observation, further thorough examinations are thus needed for their confirmation. In this study, therefore, we used a global dataset of COVID-19 cases and global climate databases and comprehensively investigated how climate parameters could contribute to the growth rate of COVID-19 cases while statistically controlling for potential confounding effects using spatial analysis. We also confirmed that the growth rate decreased with the temperature; however, the growth rate was affected by precipitation seasonality and warming velocity rather than temperature. In particular, a lower growth rate was observed for a higher precipitation seasonality and lower warming velocity. These effects were independent of population density, human life quality, and travel restrictions. The results indicate that the temperature effect is less important compared to these intrinsic climate characteristics, which might thus be useful for explaining the exceptions. However, the contributions of the climate parameters to the growth rate were moderate; rather, the contribution of travel restrictions in each country was more significant. Although our findings are preliminary owing to data-analysis limitations, they may be helpful when predicting COVID-19 transmission.

## 1. Introduction

The world-wide spreading of coronavirus disease 2019 (COVID-19) [1], an infectious disease caused by the novel coronavirus, severe acute respiratory syndrome coronavirus 2 (SARS-CoV-2 / 2019-nCoV) was firstly identified in Wuhan, China [2]. The COVID-19 epidemic has a serious impact on the public health and economy [3], the reduction of its spreading is thus a significant challenge. How climate parameters are associated with the spreading is intriguing concerning the coronavirus characterization and spreading prediction. Previous studies have suggested that temperature increase could reduce COVID-19 transmission both in China [4–6] and at the global scale [7–11]. However, a bell-shaped or quadratic relationship between the COVID-19 transmission rate and the temperature was observed, indicating that the optimal transmission temperature could be at ∼8 °C. Moreover, part of the previous studies [4–6, 8] also reported that higher humidity is also associated with a lower transmission rate of COVID-19. These results are consistent with the influenza seasonality (i.e., the fact that influenza transmission is reduced due to temperature and humidity increase) [12]. Thus, previous studies have predicted that the arrival of summer and the rainy season would reduce COVID-19 transmission.

However, more careful examinations are required to conclude such COVID-19 seasonality. As emphasized in part of the previous studies, temperature could account for a relatively modest amount of the total variation in the COVID-19 transmission rate [10]. In fact, despite the expectations, the spreading of COVID-19 has also been observed in warm and humid areas (e.g., Australia, Brazil, and Argentina, on Southern Hemisphere, in early March). This indicates that other climate parameters might also affect COVID-19 transmission. For example, influenza transmission is also influenced by several environmental parameters, such as ultraviolet (UV) radiation, wind speed, precipitation, and air pollution. [13]; moreover, it also correlated with diurnal temperature ranges [14] and urbanization (human impacts) [15]. In addition to this, changing rapid weather variability (e.g., climate seasonality and climate change) increases the risk of an influenza epidemic [16]. In general, seasonal variations in temperature, rainfall, and resource availability can exert strong pressure on infectious disease population dynamics [17]. Inspired by these results, previous studies evaluated the contributions of wind speed [8], precipitation and UV irradiation to COVID-19 transmission [9]. However, the remaining parameters have still been poorly investigated to date. In particular, the temperature might be associated with other climate parameters, it is thus necessary to control the potentially confounding effects [18–20].

To study the aforementioned subject, the application of spatial analysis might also be needed. Although spatial autocorrelations between observation areas and variables need to be evaluated when analyzing geographic data [18, 20, 21], previous studies have understudied them. It remains possible that the observed associations of COVID-19 transmission with temperature and humidity are spatial autocorrelation artefacts.

In this study, we thus aimed at conducting a more comprehensive investigation. Using global time-series data on confirmed COVID-19 cases[1] and global climate databases, we comprehensively investigated how climate parameters contribute to COVID-19 transmission on a global scale while statistically controlling for potential confounding effects using spatial analysis. Population density and quality of human life (human development index) were also considered when controlling for potential confounding effects because they might affect infectious disease transmission [15], including COVID-19 transmission [4]. Similarly, we also considered the travel restrictions because the national emergency response, including travel bans, appears to have delayed the growth and limited the size of the COVID-19 epidemic in China [22, 23].

## 2. Material and methods

### 2.1. The growth rate of COVID-19 cases

We obtained global time-series data for the period between January 22, 2020 - April 6, 2020, on the number of confirmed cases of COVID-19 [1] operated by the Johns Hopkins University Center for Systems Science and Engineering from their GitHub repository. In this repository, the global dataset and dataset of the (USA) were available. We combined these datasets after removing USA-related data from the global dataset. To estimate the COVID-19 transmission rate, many previous studies considered the measures based on the number of cases. However, it remains possible that the differences in the number of tested individuals between areas (countries) affect these measures. We thus used instead the growth rate of confirmed COVID-19 cases as a more suitable measure. The growth rate in each observation was computed using the R statistical software (version 3.6.2; www.r-project.org) and the package *incidence*(version 1.7.1) [24]; in particular, the *fit* function was used. To estimate the growth rate during the initial (exponential) phase, we used the data within 15 days (∼2 weeks) starting from the date (call *first date* hereafter) when 30 and more cases were confirmed in cumulative counts, as described previously [7]. We confirmed that similar conclusions were obtained at the different cut-off values (using the data within 30 days starting from the date when 50 and more cases were confirmed).

### 2.2. Climate parameters

We obtained climate parameters from several databases based on the observation area latitudes and longitudes available in the dataset [1]. The data extraction and calculation of climate parameters were generally based on the procedures established in our previous publications [18, 20], which could be also accessed in our GitHub repository [25].

Climatic parameters with a spatial resolution of 2.5’ were obtained from the WorldClim database (version 2.1) [26] for each observation area. In particular, we extracted the following monthly climate data according to the month of the median date in the data used for computing the growth rate: monthly mean temperature (*T*_mean_; °C), minimum temperature (*T*_min_; °C), maximum temperature (*T*_max_; °C), precipitation (mm), wind speed [ms^−1^], solar radiation (UV; kJ m^−2^day^−1^), and water vapor pressure [kPa]. Moreover, we computed monthly diurnal temperature range (i.e., *T*_max_ – *T*_min_; DTR; °C) and relative humidity based on *T*_mean_ and water vapor pressure. We also obtained the following annual climate parameters: temperature seasonality (*T*_seasonality_; standard deviation) and precipitation seasonality (*P*_seasonality_; coefficient of variation).

In order to evaluate the historical climate change, we computed warming velocity (WV) [27, 28], defined as the temporal annual mean temperature (AMT) gradient divided by the spatial AMT gradient, where the temporal gradient is defined as the difference between the current and past AMT, available in the WorldClim database, and the spatial gradient was the local slope of the current climate surface at the observation area, calculated using the function *terrain* (with the option neighbors = 4) in the R package *raster* (version 2.9.5).

### 2.4. Other related parameters

To investigate the effect of population density, we obtained 2020 population density (PD) data with a spatial resolution of 2.5’ from the *Gridded Population of the World* (version 4) [29].

To evaluate human impact, we used the human footprint (HF) scores, obtained from the *Last of the Wild Project* (version 3) [30]. The HF scores have a spatial resolution of 1 km grid cells and are defined based on human population density, human land use and infrastructure, and human access.

To evaluate the quality of human life, we used the gross domestic product (GDP) per capita and human development index (HDI), obtained from the *Gridded global datasets for Gross Domestic Product and Human Development Index over 1990-2015* [31]. HDI is defined based on life expectancy, education, and income (GDP per capita).

To evaluate the effect of travel restrictions, we manually extracted the dates when travel restrictions were imposed in each country from the Wikipedia page “*Travel restrictions related to the 2019–20 coronavirus pandemic”*[32]. The travel restrictions were classified into three categories: countries and territories implementing a global travel ban (Ban), countries implementing global quarantine measures (Qua), and non-global restrictions (NonG). When a country imposed multiple restriction types, the date when the strongest restriction was imposed was selected, where the order of the strength of travel restrictions was considered as follows: Ban > Qua > NonG. Many countries imposed travel restrictions after March 17, 2020 (see Figure S1 in our GitHub repository [25]). Thus, we considered a categorical variable (Ban) for the global travel restriction trend: 0 if the first date (see Section 2.1) is before March 17, 2020, and 1 otherwise.

### 2.3. Data analyses

The statistical analyses were based on the procedures in [18, 20]. To evaluate the contribution of each variable to the growth rate, regression analysis was performed using R. Both ordinary least-squares (OLS) regression and the spatial analysis approach were considered. The dataset and R script for data analyses, used in this study, are available in our GitHub repository [25].

For the OLS regression, full models were constructed encompassing all explanatory variables (*T*_mean_, DTR, *T*_seasonality_, wind speed, precipitation, *P*_seasonality_, UV, humidity, PD, HDI, WV, and Ban), and the best model was selected in the full model. The HF scores and GDP per capita were omitted because they were strongly correlated with PD and HDI, respectively. The best model was selected based on the sample-size-corrected version of the Akaike information criterion (AICc) values using the R package *MuMIn* (version 1.43.6). Moreover, a model-averaging approach using *MuMIn* was adopted. The averaged model was obtained in the top 95% confidence set of models. A global Moran’s test was performed to evaluate spatial autocorrelation in the regression residuals using the function *lm.morantest.exact* in the R package *spdep*, version 1.1.3.

PD and WV were log-transformed for normality. Precipitation and *P*_seasonality_ were square-root transformed for normality. *T*_mean_ was rescaled with 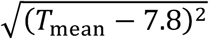 to the quadratic relationship between temperature and transmission rate of COVID-19 [5, 7, 8, 10, 11]. The quantitative variables were normalized to the same scale, with a mean of 0 and a standard deviation of 1, using the *scale* function in R before the analysis.

For spatial analysis, a spatial eigenvector mapping (SEVM) modeling approach [21, 33] was also considered to remove spatial autocorrelation in the regression residuals.

Specifically, the Moran eigenvector approach was adopted using the function *SpatialFiltering* in the R package *spatialreg* (version 1.1.5). As with the OLS regression analysis, full models were constructed, and then the best model was selected based on AICc values. The spatial filter was fixed in the model-selection procedures [33].

The contribution (i.e., non-zero estimate) of each explanatory variable to the growth rate of COVID-19 cases was considered significant when the associated *p*-value was less than 0.05.

## 3. Results and discussion

The data in 300 areas were investigated (Figure 1). The OLS regression analysis (Table 1) and spatial analysis (Table 2) showed almost similar results because the statistical significances of spatial autocorrelations were moderate in the full model (Moran’s *I* = 0.077, and the associated *p*-value = 0.021) and best model (*I* = 0.084, *p* = 0.027) of the OLS regression analysis. The full, best, and averaged models showed almost similar results in both the OLS regression analysis and spatial analysis. The details of the results are as follows.

**Table 1.** Influence of explanatory variables on the growth rate of COVID-19 cases based on the ordinary least squared regression approach. The results of the full model, best model, and averaged model are shown, respectively. The abbreviations of variables are as follows: *T*_mean_ (monthly mean temperature), DTR (monthly diurnal temperature range), *T*_seasonality_ (temperature seasonality), *P*_seasonality_ (precipitation seasonality), UV (monthly solar radiation index), WV (warming velocity), PD (population density), HDI (human development index), and Ban (travel restrictions). *R*^2^ denotes the coefficient of determination for full and best models based on the OLS regression. SE is the standard error. Values in brackets are the associated *p*-values.

| Variables | Full model |  |  | Best model |  |  | Averaged model |  |  |
| --- | --- | --- | --- | --- | --- | --- | --- | --- | --- |
| | Estimate | SE | $p$-value | Estimate | SE | $p$-value | Estimate | SE | $p$-value |
| $T_{\text{mean}}$ | −0.18 | 0.07 | 0.014 | −0.17 | 0.06 | $9.0 \times 10^{-3}$ | −0.16 | 0.07 | 0.032 |
| Humidity | −0.10 | 0.09 | 0.27 |  |  |  | −0.12 | 0.08 | 0.14 |
| DTR | 0.02 | 0.09 | 0.86 |  |  |  | 0.08 | 0.09 | 0.36 |
| $T_{\text{seasonality}}$ | −0.14 | 0.09 | 0.14 | | | | −0.13 | 0.09 | 0.15 |
| Wind speed | −0.05 | 0.07 | 0.53 |  |  |  | −0.04 | 0.07 | 0.57 |
| Precipitation | −0.03 | 0.08 | 0.73 |  |  |  | −0.01 | 0.08 | 0.90 |
| $P_{\text{seasonality}}$ | −0.30 | 0.10 | $1.3 \times 10^{-4}$ | −0.28 | 0.07 | $7.2 \times 10^{-5}$ | −0.30 | 0.08 | $9.2 \times 10^{-5}$ |
| UV | 0.13 | 0.03 | 0.18 | 0.23 | 0.07 | $5.4 \times 10^{-4}$ | 0.18 | 0.09 | 0.060 |
| WV | 0.18 | 0.07 | $9.1 \times 10^{-3}$ | 0.14 | 0.06 | 0.017 | 0.15 | 0.07 | 0.028 |
| PD | −0.06 | 0.06 | 0.27 |  |  |  | −0.06 | 0.06 | 0.35 |
| HDI | 0.24 | 0.08 | $9.1 \times 10^{-3}$ | 0.21 | 0.07 | $1.8 \times 10^{-3}$ | 0.23 | 0.08 | $1.7 \times 10^{-3}$ |
| Ban | −0.68 | 0.13 | $3.2 \times 10^{-7}$ | −0.71 | 0.12 | $1.8 \times 10^{-8}$ | −0.68 | 0.13 | $1.0 \times 10^{-7}$ |
| Moran's $I$ | 0.077 (0.021) | | | 0.084 (0.027) | | | | | |
| $R^2$ | 0.26 ($2.1 \times 10^{-13}$) | | | 0.24 ($1.2 \times 10^{-15}$) | | | | | |
| AICc | 791 |  |  | 783 |  |  |  |  |  |

**Table 2.** Influence of explanatory variables on the growth rate of COVID-19 cases based on the spatial analysis approach. The results of the full model, best model, and averaged model are shown. *R*^2^ denotes the coefficient of determination for full and best models based on the SEVM modelling. SE is the standard error. Values in brackets are the associated *p*-values. See Table 1 for description of table elements.

| Variables | Full model |  |  | Best model |  |  | Averaged model |  |  |
| --- | --- | --- | --- | --- | --- | --- | --- | --- | --- |
| | Estimate | SE | $p$-value | Estimate | SE | $p$-value | Estimate | SE | $p$-value |
| $T_{\text{mean}}$ | −0.20 | 0.07 | $4.8 \times 10^{-3}$ | −0.21 | 0.06 | $1.4 \times 10^{-3}$ | −0.20 | 0.07 | $3.1 \times 10^{-3}$ |
| Humidity | −0.03 | 0.09 | 0.73 |  |  |  | −0.05 | 0.08 | 0.55 |
| DTR | 0.07 | 0.09 | 0.47 |  |  |  | 0.09 | 0.07 | 0.21 |
| $T_{\text{seasonality}}$ | −0.01 | 0.09 | 0.89 | | | | 0.00 | 0.09 | 0.99 |
| Wind speed | −0.03 | 0.07 | 0.68 |  |  |  | −0.04 | 0.07 | 0.59 |
| Precipitation | 0.02 | 0.08 | 0.77 |  |  |  | 0.01 | 0.08 | 0.87 |
| $P_{\text{seasonality}}$ | −0.32 | 0.08 | $1.2 \times 10^{-4}$ | −0.31 | 0.07 | $1.5 \times 10^{-5}$ | −0.33 | 0.08 | $2.1 \times 10^{-5}$ |
| UV | 0.24 | 0.10 | 0.013 | 0.30 | 0.07 | $2.1 \times 10^{-5}$ | 0.27 | 0.08 | $1.0 \times 10^{-3}$ |
| WV | 0.19 | 0.07 | $3.7 \times 10^{-3}$ | 0.20 | 0.06 | $8.7 \times 10^{-4}$ | 0.19 | 0.06 | $2.8 \times 10^{-3}$ |
| PD | −0.03 | 0.06 | 0.55 |  |  |  | −0.04 | 0.06 | 0.49 |
| HDI | 0.21 | 0.08 | $9.3 \times 10^{-3}$ | 0.21 | 0.07 | $2.1 \times 10^{-3}$ | 0.21 | 0.07 | $3.1 \times 10^{-3}$ |
| Ban | −0.73 | 0.13 | $2.5 \times 10^{-8}$ | −0.72 | 0.12 | $4.8 \times 10^{-9}$ | −0.73 | 0.12 | $< 2.0 \times 10^{-16}$ |
| Moran's $I$ | −0.052 (0.51) | | | −0.054 (0.60) | | | | | |
| $R^2$ | 0.33 ($2.9 \times 10^{-16}$) | | | 0.32 ($< 2.2 \times 10^{-16}$) | | | | | |
| AICc | 773 |  |  | 763 |  |  |  |  |  |

**Figure 1.**
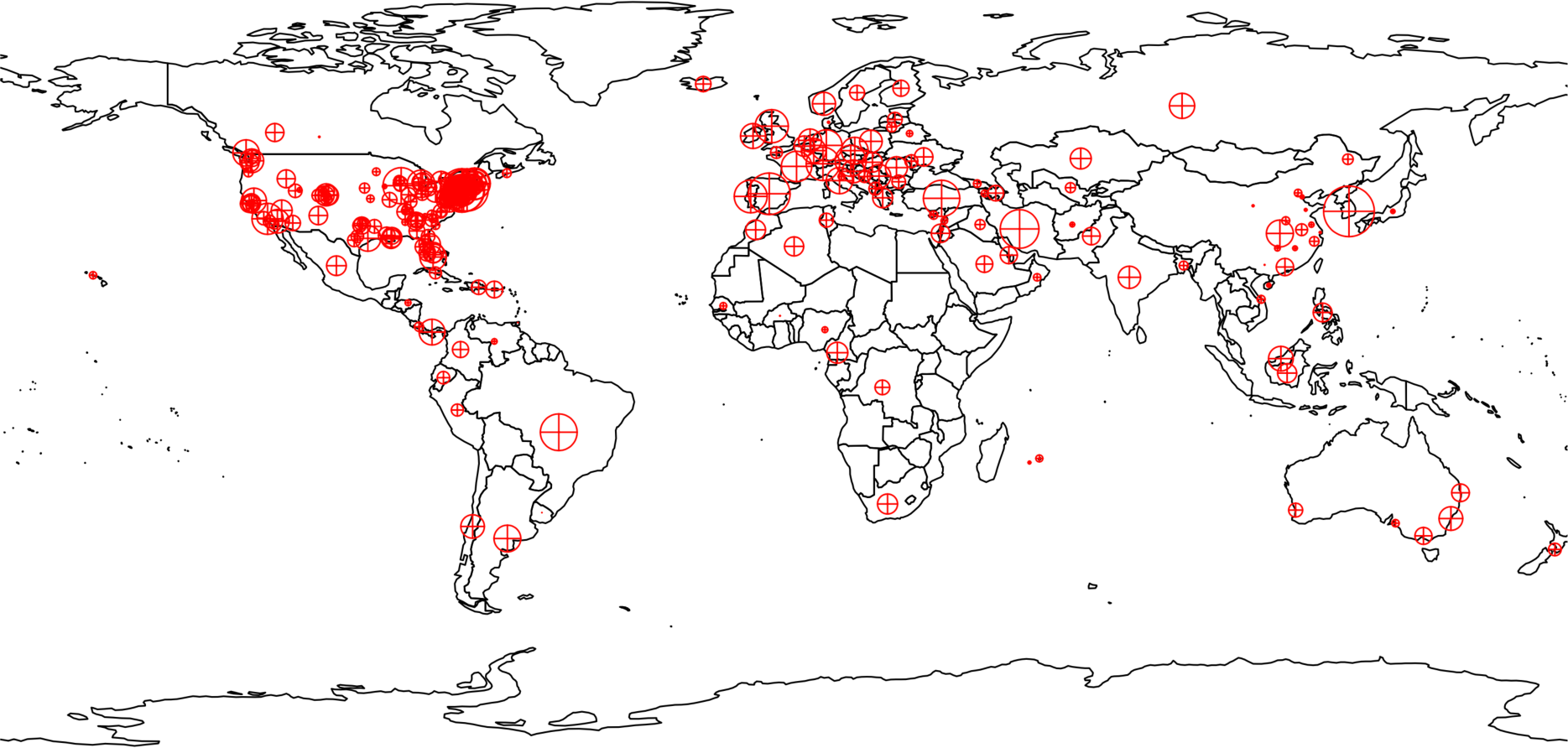
Distribution of the observation areas included in this study. Red symbols indicate the observation areas. Symbol size indicates the growth rate of COVID-19 cases.

The temperature negatively correlated with the growth rate of COVID-19 cases. This indicates that high temperature (e.g., the arrival of summer season) reduces COVID-19 transmission, consistent with several previous studies [4–11]. However, no humidity contribution was observed. This discrepancy might be due to differences in the datasets and data analyses between this study and previous studies. Previous studies (e.g., [4]), reported the association with humidity, was limited to the data on China; moreover, they used the measures based on the number of confirmed cases, although these measures may be affected by the difference of COVID-19 testing between areas. The contribution of humidity may be limited on a global scale. A similar tendency is observed in the case of influenza [13]; in particular, using specific humidity to determine transmission has a low predictive power at low- and mid-altitude sites, although humidity is believed to affect the transmission.

More importantly, however, we found that the growth rate was associated with the other parameters rather than temperature. In particular, we found that the growth rate of COVID-19 cases showed a correlation with precipitation seasonality and warming velocity. Specifically, a lower growth rate was observed during a higher precipitation seasonality and a lower warming velocity; however, the contribution of precipitation seasonality was higher than that of warming velocity, according to the estimates of the models of the OLS regression analysis and spatial analysis. The observed associations may be reasonable in the context of the effects of seasonality and changing rapid weather variability on population dynamics of infectious diseases [17]. In particular, theory and experiment have indicated that climate seasonality can alter the spread and persistence of infectious diseases and that population-level responses can range from simple annual cycles to more complex multiyear variations. Therefore, climate seasonality and historical climate change can affect infectious disease transmission. In fact, rapid weather variability played a significant role in changing the strength of the influenza epidemic in the past [16]. However, the reason why temperature seasonality did not correlate with the growth rate remains unclear. Nevertheless, these results (the contribution of precipitation seasonality, in particular) may explain the exceptions (i.e., why the spreads of COVID-19 are also observed in warm areas although previous studies suggest that high temperature reduces COVID-19 transmission). This may be because of the difference in precipitation seasonality between the observation areas. For example, the areas in Australia, Brazil, and Argentina were warm in March; however, they show low precipitation seasonality (Figure 2). Thus, the spreads might occur in these areas. Moreover, Europe and the USA might have undergone rapid spreads because they show low precipitation seasonality; on the other hand, the spread might have reached a peak relatively quickly in China because of relatively high precipitation seasonality.

**Figure 2.**
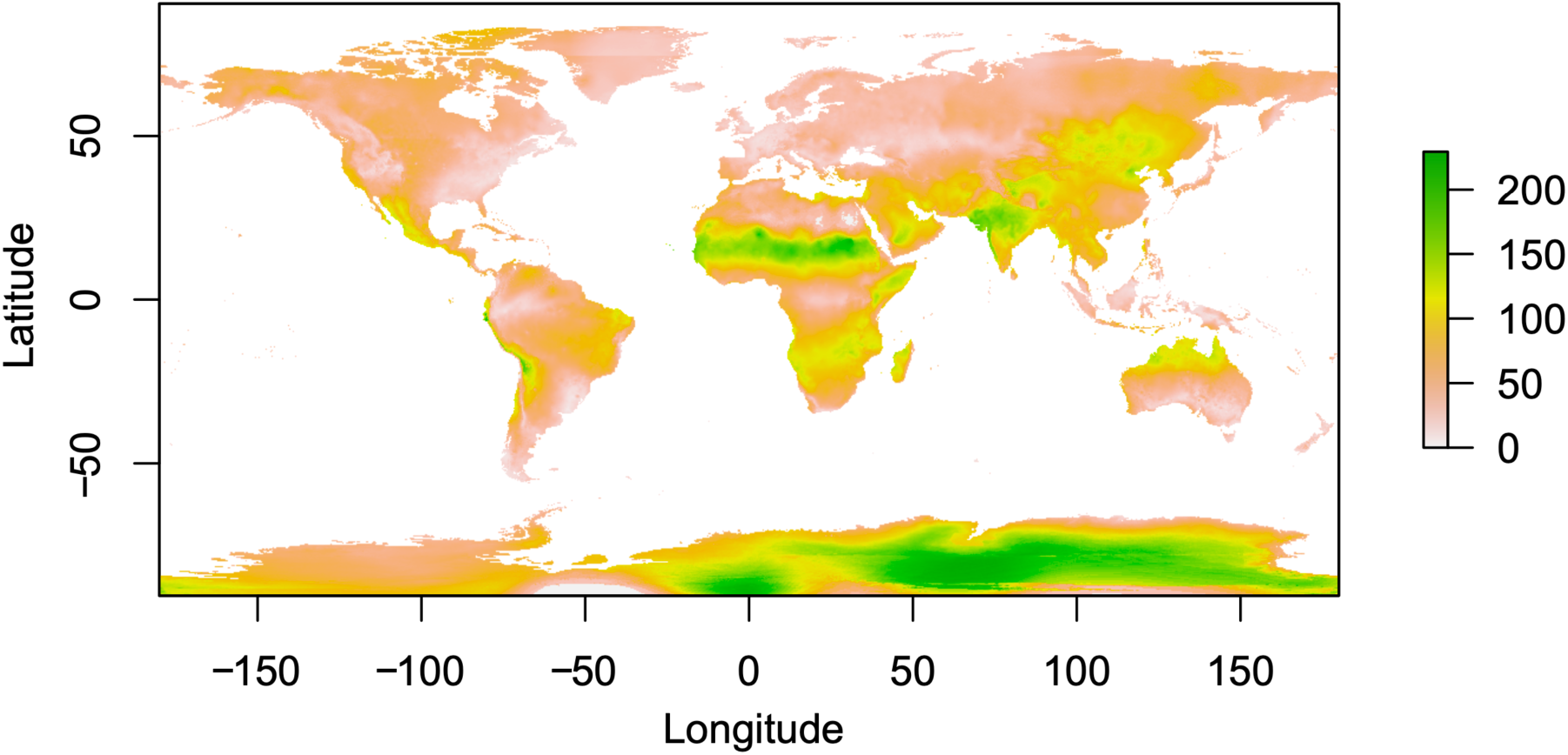
World distribution of precipitation seasonality.

The contribution of solar radiation is currently ambiguous. Solar radiation showed a positive association with the growth rate of COVID-19 cases. However, the results were less robust; in particular, the contribution was statistically significant in spatial analysis (Table 2), but not in the full and averaged models in the OLS regression (Table 1). Thus, it remains possible that the contributions partly observed in the analyses are artefacts. Assuming the positive association, the result is inconsistent with the fact that solar (UV) radiation is expected to reduce infection disease (e.g., influenza) transmission [13]. Moreover, a pairwise correlation analysis showed no association between the growth rate and solar radiation (Spearman’s rank correlation coefficient *r* = –0.06, *p* = 0.31).

The contributions of wind speed and precipitation were also limited. This is inconsistent with previous studies [8, 9]; however, statistical significances were not evaluated in these studies. This discrepancy might be due to differences in the data analyses between this study and previous studies. In particular, previous studies used the measures based on the number of confirmed cases; however, these measures may be affected by the difference of COVID-19 testing between areas. Hence, further examinations may be needed, given the importance of these climate parameters in infectious disease transmission [13, 17].

Non-climate parameters were also associated with the growth rate of COVID-19. According to the estimates of the models of the OLS regression analysis and spatial analysis, the contribution of travel restrictions was most significant than those of the climate parameters; in particular, travel restrictions showed a negative association with the growth rate. This result may be an extension of the result that the reduction of COVID-19 transmission due to interventions, including travel restrictions, in China [22, 23]. Our result implies that the travel restrictions in each country contributed to reducing COVID-19 transmission on a global scale.

The quality of human life (HDI) showed a positive association with the growth rate of COVID-19. This may be because HDI reflects life expectancy (i.e., areas with a higher HDI tend to have more older individuals because of a higher quality of human life). COVID-19 has the age specificity of cases and attack rates [34]; in particular, the epidemic risks of disease given exposure are likely to be the highest among adults aged from 50-69 years. Thus, the growth rate is expected to increase with HDI.

## 4. Conclusions

Intrigued by the question why COVID-19 transmission is observed in warm areas despites previous expectations of COVID-19 transmission reduction at high temperatures, we comprehensively investigated how several climate parameters are associated with the growth rate of COVID-19 cases and found that it was affected by precipitation seasonality and warming velocity rather than temperature. The effects were independent of population density, quality of human life, and travel restrictions. Our findings must necessarily be considered preliminary due to several limitations; in particular, it remains possible that the observed association is indirect. However, they may enhance our understanding of the COVID-19 transmission. As previous studies mentioned, high temperatures might reduce COVID-19 transmission. However, the effects may be restricted by intrinsic climate characteristics, such as precipitation seasonality and warming velocity. Moreover, the contributions of climate parameters to the growth rate of COVID-19 cases were moderate, while those of national emergency responses (i.e., travel restrictions) were more significant. Thus, slowing down the spread of COVID-19 due to the arrival of the summer season might not be expected. Instead, global collaborative interventions might be necessary to halt the epidemic outbreak.

## Data Availability

The datasets generated and analyzed in the current study are available in the GitHub repository: https://github.com/kztakemoto/covid19climate. The relevant R codes can be also found in the GitHub repository.

## Ethics

This study required no ethical permission.

## Authors’ contributions

KT conceived and designed the study. KC and KT prepared the data and performed data analysis, interpreted the results, and wrote the manuscript. Both authors gave their final approval for publication.

## Acknowledgements

The authors would like to thank Editage (www.editage.com) for English language editing.

